# Challenges of visibility and inclusion in Long COVID: a population-based interview study

**DOI:** 10.1101/2025.04.08.25325457

**Authors:** Emily Cooper, Adam Lound, Kathryn Jones, Jane Bruton, Sophie Day, Christina J Atchison, Caroline Eccles, Alex Piper, Graham S. Cooke, Marc Chadeau-Hyam, Paul Elliott, Helen Ward

**Author notes:** Corresponding author: Helen Ward. Email addresses. **Data availability** All relevant data are within the manuscript of this qualitative study. Full transcripts/data cannot be shared publicly because of the sensitive and potentially identifiable nature of the qualitative data we collected, and the nature of the consent obtained from participants which ensured confidentiality and anonymity. Data access requests for elements of raw data may be sent to the REACT Data Access Committee (contact via). **Ethics** The REACT and REACT-LC Studies hold ethical approval from South-Central Berkshire B Research Ethics Committee (IRAS IDs: 298404, 259978, 283787, 298724). All participants provided informed consent to take part in the study. **Patient consent statement** All participants provided informed consent. **Permission to reproduce material from other sources** N/A. **Clinical trial registration** N/A.

## Abstract

**Introduction:** People with Long Covid report a wide range of symptoms and inconsistent responses when seeking clinical diagnosis and support. Much of the qualitative research on Long Covid has been based on people attending specialist clinical services or who have accessed support groups. We describe the varied ways in which a diverse group of people in a community sample experience, recognise and manage persistent symptoms following COVID-19.

**Methods:** Qualitative interview study nested within the large, community-based REal-time Assessment of Community Transmission (REACT) study in England. Participants reporting persistent symptoms following COVID-19 were asked for consent to be contacted about a follow-up interview. We then purposively sampled by age, gender, ethnicity and symptom severity and conducted 60 interviews. Analysis was carried out using a reflexive thematic analysis approach.

**Results:** Participants were an ethnically diverse group aged between 18 and 80 years who reported symptoms following COVID-19 for between a few months and more than two years. Many had not accessed clinical care or specific Long Covid support, and some did not identify with the category of long covid, rendering their experiences largely invisible. Participants highlighted the ways in which they self-manage symptoms within this context, and the varied burden of coping with ongoing health problems.

**Conclusion:** This diverse sample of people with Long Covid report a range of challenges managing this emerging and contested condition, with uncertainty affecting their own understanding and the validation they receive from professionals, family and friends. These challenges intersect with others such as racism, and are compounded by a lack of specific resources for Long Covid as well as over-stretched health services in the UK. Nevertheless, people show resilience in their self-management, seeking information and support from a range of sources.

**Patient or Public Contribution:** Study design, analysis and outputs informed by Patient Advisory Group.

## Introduction

Long Covid (also known as Post-COVID condition or post-COVID syndrome) is a significant public health challenge (1,2) with diagnostic uncertainty and gaps in knowledge. While biomedical research and clinical trials into potential mechanisms and treatments are ongoing, patients and clinicians are left with inadequate guidance and support (3,4).

In 2020, Long Covid support groups were established to bring people together to share experience, provide support and advocate for research and treatment. These groups continue to play a critical role helping patients, carrying out research, and informing clinical services that developed in a number of countries (5,6). However, five years later people with persistent symptoms following COVID-19 are often still unable to access clinical care or support (7).

Most published evidence on the experiences of people with Long Covid comes from studies recruiting from support groups or clinical services (7,8). These studies therefore reflect experiences of those who have already navigated their way to some kind of support, including a predominance of women and people of white ethnicity. It is unclear whether their experiences are shared with a more demographically diverse group of participants who may be managing their symptoms outside of Long Covid support groups or clinical services. These latter groups are likely to be relatively hidden from routine data and research accounts of Long Covid.

In this paper we report on interviews from a diverse sample of people who reported persistent symptoms following COVID-19 as part of the follow-up of people from the REal-time Assessment of Community Transmission (REACT) population-based study (9,10). In a pilot study we identified significant variation in symptom experience and differences in awareness or perceived applicability of the term ‘Long Covid’, and highlighted how lack of recognition could prevent people from seeking care (11). Other studies have focused on barriers to accessing treatment and care (12). Here we explore the varied ways in which people experience, recognise and manage persistent symptoms following COVID-19.

## Materials and methods

### Sample and recruitment

This interview study is nested within the large REACT-Long Covid project (REACT-LC), itself based on follow-up of participants in REACT (for detailed methods of the REACT studies see 9,10,13). Briefly, between 2020 and 2022 we invited random samples of the population of England to perform nose and throat swabs (REACT-1) or finger-prick blood tests (REACT-2) to estimate the prevalence of the SARS-CoV-2 virus and IgG antibodies respectively; over 3 million adults took part, a majority of whom consented to being contacted for further research. In 2022, over 270,000 people completed a follow-up questionnaire on health and wellbeing as part of the REACT-LC study (13): a sub-sample of 10,500 adults with a history of SARS-CoV-2 infection had a clinical examination and biological samples taken at a REACT-LC assessment clinic.

Eligibility for this interview study was being a REACT participant who had reported persistent symptoms for 12 or more weeks following symptomatic COVID-19, and who either (a) attended one of the REACT LC assessment clinics or (b) completed the REACT-LC Health and Wellbeing survey and consented to be approached for interview. We purposively sampled by age, gender, ethnicity and symptom severity. We aimed for 60 interviews to ensure an inclusive sample with a range of experiences.

For those recruited via the REACT-LC assessment clinics, invitation emails with a participant information sheet and link to an online consent form were sent out to 188 people who met our inclusion criteria, interviews were arranged at mutually convenient times until we had completed 27 interviews.

Further participants were recruited following the first two waves of the survey fieldwork; 18,827 people met our inclusion criteria, of whom 15,634 (83.0%) indicated willingness to be recontacted for interview. These were grouped by our sampling criteria, and 91 were contacted; interviews were arranged at mutually convenient times until 33 were completed. At total of 60 participants were recruited and interviewed across both routes and no participants dropped out.

### Topic Guide

The topic guide was initially developed for a pilot interview study (14) informed by existing literature and together with the REACT public advisory committee. It focused on experience of acute and persistent symptoms, the impact of those symptoms, plus diagnosis and management. After the pilot study, the topic guide was reviewed, and additional sections added on pre-pandemic life, self-management and recovery. The semi-structured style of the guide provided questions on broad topics for discussion using prompts and probes to elucidate more detail from participants. The topic guide is available in S1 Appendix 1.

#### Data collection

Interviews were conducted on-line using MS Teams or Zoom by one of two interviewers (EC and AL) and audio recorded with permission. Interviews lasted around an hour. Notes were also taken to aid recall during analysis. Recordings were transcribed verbatim by a transcription service. All data were encrypted and stored in a secure folder on Imperial College servers, which could only be accessed by members of the research team. The data were managed using NVivo 13 R1 (released 2020). Interview responses were linked to survey responses by a unique study number. Contact details were made available to interviewers via a secure data enclave for the purposes of arranging the interview only. Demographic and interview data were kept separate from recruitment data and linked only by a study number.

#### Analysis

Analysis was carried out using a reflexive thematic analysis approach (15). This first wave of inductive coding was carried out by EC, KJ, JB and AL. Codes were reviewed and compared by EC and AL and a second wave of coding was conducted to confirm or clarify existing codes and capture any new ideas or descriptions that had been missed. A mix of semantic and latent codes were used, allowing for an exploration of both the participant’s explicit understandings and the possible underlying meaning behind their narratives. Once the codes were agreed the research team began to look for patterns in the data and ‘central organising concepts’ (16). Themes were explored and developed as either descriptive themes, reflecting topics explored in the interview guide, or analytic themes which expressed new or emergent ideas.

### Reflexivity statement

EC is a white female qualitative researcher with a background in anthropology and health research. AL is a white male physiotherapist and qualitative researcher. HW is a white female public health clinician and senior researcher leading the REACT programme who works closely with patient advocates and Long Covid charities.

### Public Involvement

The REACT-LC advisory group were involved throughout the pilot and main study. This included input into the topic guide and study materials, discussion of preliminary findings and development of outputs. The group were pivotal in the development of the research questions for this interview study (17).

### Governance

This study received ethical approval from the South-Central Berkshire B Research Ethics Committee (IRAS ID: 298404, REC Ref 21/SC/0134) and all participants provided informed consent to be interviewed

## Results

### Characteristics of the sample

The 60 participants were aged between 19 and 80, 23 identified as men, including one transgender man, and 37 as women. By broad categories of ethnicity, 29 participants were white, 10 Black, 9 Asian and 12 Mixed or other. A majority (33) lived in London or the South East, with others from across England. Most reported symptomatic COVID-19 in 2020 (n=46), 3 in late 2019 (IgG antibody positive in REACT-2), 10 in 2021 or 2022. One person described symptoms of COVID-19 on multiple dates.

Most (n=53) felt that they had not yet recovered from COVID-19 symptoms, while seven reported feeling recovered (one between four and six months, three between six months and a year, three between one to two years). For those who were still experiencing symptoms, the duration varied from three to five months (n=3), six to 11 months (n=5), 12 to 23 months (n=24) and 2 or more years (n=21). Almost all reported some degree of improvement, either over time or during phases between flare ups; the fluctuating nature of symptoms made recovery status hard to quantify. In terms of recovery trajectories, 41 described their recovery as gradual and seven reported fluctuating improvements. Three reported sudden improvement; one after having a COVID-19 vaccine, one after another episode of COVID-19, and one with no specific trigger. Two reported recovering after receiving treatment.

Fatigue was the most common persistent symptom, mentioned by 42 of the 60, with breathing issues and cognitive symptoms mentioned by 23 and 24 of participants respectively. Loss of taste and/or smell were reported by 16 participants.

### Severity and impact of symptoms

Based on the interviews we broadly categorised the severity of people’s symptoms in relation to their impact on usual activities with 17 being ‘severe’, meaning they were unable to participate in usual activities and 43 ‘less severe’ as they were able to do usual activities, but with exertion often leading to worsening symptoms. When asked directly, two thirds of the participants (n=39) described themselves as having Long Covid, 12 did not, and nine reported being ambivalent about using the term. All but one of those who did not use, or were ambivalent, about the term fell into the less severe category. Of the seven participants who had recovered at the time of interview, four had considered themselves to have had Long Covid and three did not.

### Experience of healthcare

Almost half of our participants (n=27) had not consulted a clinician about their persistent symptoms, and of those who did, some only mentioned their symptoms during an appointment for a different or pre-existing health issue. This healthcare seeking was associated with severity, with all of those categorised as severe and under half of those who were less severe seeking care from clinical services. Reasons for not seeking help varied. Some did not attribute their ongoing symptoms to COVID-19, despite describing them as persisting since initial infection, preferring alternative explanations such as ageing, menopause or deconditioning. Others felt that their symptoms were manageable without treatment, did not want to burden the health service, and some suggested seeking treatment was futile since there was no available treatment.

‘*It’s something that I’ve got to live with now and it’s not going to get better. It’s not disabling me and it’s clearly not curable. I’m not going to go to the doctor because I can’t taste*.*’* (AL3, White, F, 50-64, Less Severe, 4-5 months, Recovered) This participant considered themselves fully recovered despite still having ageusia (loss of taste).

Those who sought treatment reported different experiences. While some felt supported, others described frustrating attempts to access care, a lack of continuity and long waiting times for specialist review or therapy. This led some to seek private clinical care. Several described being met with disbelief and a lack of sympathy from clinicians; while they had hoped to receive clarity they were met with uncertainty. Some participants described feeling *‘let down’* by General Practitioners (GPs, family doctors in the UK) who did not take symptoms seriously or who dismissed the idea that the symptoms were related to COVID-19. Treatment suggestions were often felt to be inappropriate such as recommending exercise or homeopathy. However, others reported sympathetic GPs who provided advice or who were open to referring on for specialist review or investigations related to specific symptoms.

Experiences of specialist Long Covid clinics also varied. Participants who were positive appreciated the *‘holistic’* and *‘multidisciplinary’* approach. Specialists provided specific strategies for managing fatigue or breathlessness and recommended online courses and groups which allowed sharing of experiences with others. Some people described receiving personalised treatment, indicating they had learnt new skills in relation to managing fatigue, for example. Others did not feel that their existing self-management strategies had been enhanced, and one participant described their care as *‘appalling’* and said that they ‘*… haven’t heard of anyone saying that they’ve got professional help for Long Covid and it’s helpful and a game changer’. (AL22, Asian F, 50-64, Less Severe, 2 years +, Not fully recovered)* People from ethnic minority groups reported barriers to accessing and navigating healthcare services for Long Covid, with some perceiving the healthcare system as *‘institutionally racist’*, leading to disengagement and resentment:

*‘…because of my slightly darker complexion, there’s just this… I don’t know, a perception that we are tougher and able to cope with pain more than what white people’* (AL17, Black – Other, F 30-49, Less Severe, 1 year+, Not fully recovered)

There were also concerns about the disproportionate impact of COVID-19 on ethnic minority groups: *‘…on the TV every day, you just see all these photos of all these people, ethnic minority groups, sort of, dark skin, have died*.*’* (EC15, Black – Caribbean, F, 65+, Less Severe, 1 year+, Not fully recovered)

Despite awareness of the increased risks, participants were disappointed that ethnicity and comorbidities did not appear to influence how they were treated when seeking help. *‘Nobody considered the ethnic profile, they considered the clinical profile’* and were concerned that the healthcare system tended to *‘focus on the majority to the huge detriment of the minority’*. (AL27, Mixed (White and Asian), M, 50-64, Less Severe, 1 year +, Not fully recovered)

The general lack of recognition of Long Covid by clinicians was seen to be exacerbated by a wider invisibility related to gender and ethnicity:

*‘I have felt that, as an Asian woman going to doctors and things, I don’t feel heard always. And I do find, sometimes when I take my husband with me places, I get heard more*.*’* (AL18, Asian, F, 50-64, Severe, 1 year+, Not fully recovered).

### Support groups for Long Covid

A small number of participants had joined online Long Covid support groups and found opportunities to share experiences, validate symptoms, and obtain advice on self-management and navigating clinical pathways. Engagement with support groups was influenced by various factors including symptom severity, age and gender. Some people felt they did not require support, while others did not consider the groups catered for people like them:

*‘Because everything I’ve come across feels very geared for people who I feel are more poorly than I am. I feel as if there isn’t a sort of, ‘not extreme’ support group*.*’* (EC25, Black – African, F, 30-49, Less Severe, 2 years+, Not fully recovered)

There were clearly mixed feelings about support groups from those who had joined, with some finding repeated exposure to other people’s challenging experiences *‘depressing’*, others found online interactions *‘exhausting’*. Another described frustration *‘because they were getting treatments, people were seeing neurologists, they were seeing cardiologists, I was thinking, “How did you get access to that?”‘* (AL18, Asian, Female, 50-64, Severe, 1 year+, Not fully recovered). Some did not use social media or were unaware that groups had been established.

Others felt they had enough existing support describing friends, family, employers and spiritual communities who were listening, *‘checking in’* and attempting to understand what they were going through.

### Approaches to self-management

Participants reported a variety of approaches to managing symptoms and of sources of advice. Those with fatigue and breathlessness described a range of complex strategies to avoid ‘*running out of energy’* and to *‘get things done’*. Some parcelled activities into chunks, rationing how many times to go up and down the stairs in a day, for example, to avoid ‘*waste*’. They took a proactive approach to planning rest and activity with consequences in mind. ‘*If I know I’ve got something on then I won’t do anything the day before, even if that’s just meeting a friend or going to see somebody or something*.’ (EC10, White British, F, 50-64, Severe, 2 years+, Not fully recovered)

People carefully managed their rest, for example planning day-time sleep following a particular activity:

*‘I couldn’t do a day’s work on a laptop …. I taught a 30-minute lesson in the morning. I’d go and have a 2-hour sleep, 3-hour sleep sometimes, come back and just check that the kids had done their work and then that would be it, log off for the* day’. (EC13, White British, F, 30-49, Severe, 1 year+, Fully recovered).

Where participants reported pushing themselves, it was a process of trial and error, and it was evident there were contradictory consequences. While some were able to achieve their goals, this was not the case for all. Others reported negative consequences, describing how they would ‘*pay for it’* later. Some found themselves in a pattern of pushing through daily activities to the point of exhaustion or developing new or worsening symptoms.

Some participants reported needing to stop virtually everything, including work, to reduce stress, stimulations, and other demands to take time to focus their energies on their recovery. A common reflection was the need to ‘*allow’* themselves time and space to be ill and respond to their own needs. Others tried to push though their symptoms to get back to normal or have ‘*some sort of life’*. For some, this meant putting in additional effort to compensate for the deficits caused by their symptoms. Attempts to push themselves provided participants with a learning experience as they came to understand how their energy levels responded to activity.

Cognitive symptoms also had to be actively managed by using lists and notes, or setting alarms to remind them to do things. Others made plans and set targets to get through daily tasks.

### Finding ways to improve and treat symptoms

Beyond responding to symptoms through managing their capacity, participants sought out a range of ways to improve or treat symptoms dependent on the resources and networks available to them. Making improvements to general health and well-being was one-way participants saw they could potentially help or mitigate symptoms or prevent more appearing and facilitate own recovery. One participant described how getting fit could produce a positive effect:

‘*Sometimes I struggle in the gym and whatever, but I’m determined to, I don’t know, not let it win. (Laughter) But I know that this is massively minor compared to a lot of people who wouldn’t even be able to dream of getting on a bike’*. (EC12 White, F, 30-49, Less severe, 2 years+, Not fully recovered)

In the context of incomplete medical understanding of Long Covid, participants demonstrated a magpie-like approach, seeking answers from a multitude of sources including family, friends, social media and charities. While some displayed trepidation about trying remedies that may not have been ‘*proven*’ or without knowing *‘what’s actually in it’*, others were influenced by people they met outside a clinical setting but who they perceived to have specialist knowledge. Others sought advice from anyone ‘*who knows more than they do*’, including ‘*someone in Holland and Barrett’ [a health shop]*.

Several participants sought the help of complimentary practitioners including chiropractors, acupuncturists, massage therapists, reiki instructors, emotional freedom therapists and naturopaths. These complimentary approaches were valued for the relative ease of access, empathy and continuity of care, and considered useful for general wellbeing, as well as specific symptoms such as headaches and mental health.

### Wider support and new models of care

Participants were acutely aware of the pressures on the healthcare system and in particular primary care. Many suggested that there needed to be a wider menu of options to guide self-management that could be accessed without having to further burden these services. Some suggested services outside of a clinical pathway such as helplines.

Participants wanted to gain the knowledge and skills for self-management but were concerned that there was *‘so much disinformation, misinformation’* (EC31, Mixed, M, 30-49, Severe, 1 year+, Not fully recovered) around Long Covid and suggested the development of a *‘central information basket’* (AL19) from a trusted source. Current online NHS resources were seen as insufficient and perceived to offer only generic lifestyle advice. Although some described finding forums useful for sharing their experiences, they wanted up-to-date, evidence-based information on Long Covid.

In addition, participants felt there needed to be significant changes to ensure the NHS better served those living with Long Covid. Some felt the condition could not be managed through the current primary care model and that it should be broadened to include *‘an expanded GP service that can actually handle this’* (EC3, Any other White Background, M, 30-49, Less Severe, 6 months-1 year, Fully recovered).

Some participants specifically sought out mental health support as they felt medical care they received lacked empathy and focussed predominantly on physical symptoms. Therapy was used to help manage anxiety (generalised or specifically related to fear of re-infection and progression) or to help them to cope.

Some described the need for support from those around them and spoke positively of support such as partners taking on the *‘lion’s share’* of housework or childcare or shifting into a caring role *–’my partner is the only reason I eat’*. Some employers actively supported participants to manage work around symptoms, for example, flexible hours, working from home or adapting the work environment. One participant suggested their employer’s supportive ethos made a *‘huge difference’* to their ability to continue working despite living with persistent symptoms.

Most participants recognised the need for greater awareness of Long Covid amongst the public, employers and clinicians and suggested TV, radio and social media campaigns as well as information in healthcare settings such as posters in GP practices. A key part of awareness raising would be to provide a more nuanced understanding of the many ways in which Long Covid presents and impacts people to provide validation and counteract the doubt and dismissal faced in relation to symptoms: ‘*This is a definite thing. It’s not just something that’s made up*.*’* (E31, Mixed, M, 30-49, Severe, 1 year+, Not fully recovered).

Several identified the need for more clinical and general awareness of those with less severe symptoms, who were less likely to seek help, but who could be made aware of Long Covid and the support available to them.

## Discussion

This qualitative study of Long Covid addresses a gap identified by a systematic review and meta-synthesis in 2023 which noted that ‘more representative research is needed to understand Long Covid-related experiences from diverse communities and populations’ (18). By using this community-derived sample and applying a broad definition of Long Covid, namely symptoms for 12 or more weeks after COVID-19 (19), we include the experiences of a diverse group.

### Recognition and access to care

Of the 60 people we interviewed, only half had accessed clinical services for their persistent symptoms, and few had engaged with Long Covid support groups. They often had less severe, but still challenging, symptoms, and their experiences remain largely hidden both from routine data and from research on clinical experiences of the emerging category of Long Covid. One third did not apply the term to themselves, rendering them invisible (20,21). This lack of identification with the label and gaps in diagnosis of the condition reflect the ongoing uncertainty about symptom attribution where many symptoms are non-specific and common, leading some people, including clinicians, to offer alternative casual explanations (11).

Participants described validation of their symptom experience as a starting point for receiving care and support, but many found that Long Covid was not recognised or accepted by families, employers or clinicians. In this respect Long Covid aligns with contested conditions such as Myalgic Encephalomyelitis or Chronic Fatigue Syndrome (ME/CFS) and Fibromyalgia in which the lack of a diagnostic test and multiple uncertainties associated with the condition undermine its’ legitimacy (22,23). Dumit characterises contested conditions as ‘illnesses you have to fight to get’, highlighting the stigma and struggle for legitimacy patients face in addition to managing persistent symptoms (24).

Those who sought support described many difficulties in accessing appropriate treatments and care. Some reported the need for more empathetic care that considered the challenges of managing fatigue and the psychological effects of symptoms. Others felt clinicians saw symptoms as *only* psychological, leading them to discount physiological causes and fail to further investigate symptoms.

The uncertainty about Long Covid felt by some participants and their clinicians clearly limits access to healthcare and support, as does the perception that the definition only applies to people with significant disability. Many participants did not feel that their symptoms were severe enough to warrant engagement with support groups or clinical services. Some suggested they would feel like an imposter among people with more severe symptoms. We also identified negative perceptions relating to the current state of the healthcare system and pessimism about treatment options for Long Covid.

We found that demographic factors including ethnicity and gender influenced treatment seeking behaviour and healthcare experiences in common with a well-established picture of health inequalities in the UK (further exacerbated by the COVID-19 pandemic) as well as underrepresentation of vulnerable groups in research (7,25–27). There is a need to improve representation in Long Covid research to help understand the challenges faced by those from different demographic backgrounds.

### The work of self-managing

Participants reported significant and long-term impacts of symptoms on their lives. We show that these challenges are not confined to people accessing online support groups or clinical services. We describe the extent of the work patients undertake to cope with symptoms. We identified how people used trial and error to find solutions and do what they could to improve their health and wellbeing through diet, exercise or supplements. The responses of participants appeared to be attempts to claim power and autonomy by finding their own solutions. Participants sought additional input through using networks and community-based knowledge to treat themselves.

These actions are akin to the burden of ‘patient work’ described in many long-term conditions. Corbin and Straus have identified three main categories of patient work which are often invisible to others described as ‘illness work’, which includes the daily activities such as managing symptoms and taking medication; ‘everyday life work’, which relates to interactions with family and health professionals, as well as activities of daily living; and ‘biography work’, referring to coming to terms with illness (28,29). Our findings provide an insight into the often-invisible patient work required to manage Long Covid.

The ongoing labour of navigating a new way of being when managing an illness has also been identified by Bury’s theory of biographical disruption, he states ‘The realisation that medical knowledge is incomplete, and that treatment is based on practical trial and error, throws individuals back on their own stock of knowledge and biographical experience. The search for a more comprehensive level of explanation, a more certain basis of coping with the illness is often a long and profound one.’ (30). Our findings show that in the context of Long Covid work, participants rely heavily on their own knowledge and are engaged in a prolonged search for answers.

### The spectrum of Long Covid support: from validation to specialist care

Participants suggested multiple avenues of support that might help them with the work of managing persistent symptoms. In the face of multiple uncertainties and overwhelming symptoms many described the fundamental need to have their experience validated and for those around them to treat them with empathy. This kind of social support can be encouraged by providing information for family and friends on what it is like to live with Long Covid and what people need. In their review of the impact of family behaviours and communication patterns on chronic illness outcomes Rosland et al (31) found that family encouragement of self-reliance, autonomy, and personal achievement for the person with illness was associated with improved illness outcomes in a number of chronic conditions. Viewing these results in the context of our findings on self-management in Long Covid, reinforces the call for nuanced support that helps individuals maintain their independence and control whilst helping them make up the deficits in their capacity.

Similarly, participants suggested employers make appropriate adjustments to enable people with Long Covid to continue working, maintaining autonomy and self-reliance. Long Covid is now recognised as a disabling illness, although not all who experience symptoms meet criteria of disability as defined by the Equality Act 2010, due to differing severity and fluctuating nature of symptoms. As a result, some individuals with persistent symptoms experience stigma and discrimination at work (32).

Participants in our sample shared experiences of being dismissed, quitting their job or retiring early. While some participants were able to continue to work, they often faced debilitating consequences in the evenings or weekends. Participants whose employers actively supported them to manage workdays around symptoms found this enabled them to continue working, showing the value of improved sickness policies and flexible working arrangements. Given the varying attribution of persistent symptoms to COVID-19, the provision of these arrangements needs to be considered regardless of whether someone has an official diagnosis of Long Covid.

Participants in the study articulated a need for robust information and clarity around self-management to help them better help themselves. For some this could be delivered through a wider menu of options accessed outside of clinical pathways. Participants wanted the knowledge and skills to be made available from a trusted source to help them self-manage their condition as activated patients (33)

However, simply providing information is not enough. Empowering individuals requires patients and clinicians to work together as equals through shared decision-making and goal setting (34). By engaging with this mutual commitment, patients can improve clinician knowledge by sharing lived experience and in turn, clinicians can share control of symptom management by helping patients with problem solving skills (33). The Kings Fund 2018 report on ‘shared responsibility for health’ (25) highlights the need for support to be individually tailored to meet patient need, preferences, capability and level of activation. This requires patients and clinicians to develop a shared understanding of the nature and impact of persistent symptoms and to adapt management strategies based on patient capacity.

Our findings echo those from qualitative research from early in the pandemic which highlight that services are still not adequately meeting the needs of those living with Long Covid. We add to this an awareness of the expertise that Long Covid patients have in coping with, and seeking treatments for, their symptoms and the need for wider support to help them manage more effectively.

### Strengths and limitations

We worked closely with our REACT Long Covid public advisory group to ensure that the study was underpinned by lived experience and answered questions relevant to those with Long Covid. The study was informed by our pilot study which led to changes in our sampling approach, topic guide and analysis. We recruited via the representative REACT-LC study sample, so we were able to explore the views of those who had not joined Long Covid support groups or accessed clinical services. We purposively sampled to ensure diversity of experience and demographic background, over sampling for underrepresented groups. This enabled it us to include a wider variety of perspectives. Our analysis was strengthened by carrying out multiple waves of coding. We took a pragmatic approach to analysis to develop both descriptive themes which captured the variation in experience and explanatory themes which identified common patterns across experiences (16).

We carried out interviews online which meant those with persistent symptoms were able to participate more easily than having to travel or meet face to face. We acknowledge the sample does not contain those who are digitally excluded or those with the most severe symptoms, who would be unable to participate in an interview, The study is not longitudinal so was not able to map change over time. We did not interview the networks of those with Long Covid such as families, clinicians, carers or employers so have not been able to triangulate the data on experiences of self-management.

## Conclusion

Our study provides insights from a diverse sample on the experiences of managing Long Covid. We have identified the challenges around self-identification and inclusion which prevent participants from accessing Long Covid treatment and support. We have highlighted the significant amount of work that participants do to self-manage their symptoms. Finally, we have elucidated the spectrum of help and support that participants called for to help them manage their symptom.

While research into the biological causes and possible treatments of Long Covid is ongoing, there is still a lot that can be done to help patients manage in the absence of a treatment or cure. Crucially, the extensive patient work undertaken to self-manage symptoms needs to be made visible and should be a key consideration for those making decisions about support and symptom management provisions for people with Long Covid. We hope these findings will serve as a useful reference to help patients communicate their needs and enable employers, carers and clinicians to better target support and clinical services.

## Supporting information

Topic guide for interviews

## Data Availability

All relevant data are within the manuscript of this qualitative study. Full transcripts/data cannot be shared publicly because of the sensitive and potentially identifiable nature of the qualitative data we collected, and the nature of the consent obtained from participants which ensured confidentiality and anonymity. Data access requests for elements of raw data may be sent to the REACT Data Access Committee (contact via).

## Acknowledgements

Thank you to all the participants who agreed to share their experiences with us. We gratefully acknowledge the support provided by our PAG members and Graham Blakoe, Rob Elliot and Andy Heard.

## Supporting information

**Appendix 1. Topic Guide**

## Author contributions

**Conceptualization:** EC, AL, CJA, PE, HW

**Data Curation:** EC, AL

**Formal Analysis:** EC, AL

**Funding Acquisition:** PE, HW, GC

**Investigation:** EC, AL

**Methodology:** EC, Al, PE, HW

**Project Administration:** EC, AL, PE, HW

**Resources:** PE, HW

**Supervision:** CJA, HW

**Validation:** CJA, HW

**Writing – Original Draft Preparation:** EC, AL, HW

**Writing – Review & Editing:** EC, AL, KJ, CJA, JB, PE, HW

## References

1. Callard F, Perego E. How and why patients made Long Covid. Social Science & Medicine. 2021 Jan;268:113426.

2. Shah W, Hillman T, Playford ED, Hishmeh L. Managing the long term effects of covid-19: summary of NICE, SIGN, and RCGP rapid guideline. BMJ. 2021 Jan 22; n136.

3. Brightling CE, Evans RA, Singapuri A, Smith N, Wain LV, Brightling CE, et al. Long COVID research: an update from the PHOSP-COVID Scientific Summit. The Lancet Respiratory Medicine. 2023 Nov 1;11(11):e93–4.

4. Horwitz LI, Thaweethai T, Brosnahan SB, Cicek MS, Fitzgerald ML, Goldman JD, et al. Researching COVID to Enhance Recovery (RECOVER) adult study protocol: Rationale, objectives, and design. PLoS One. 2023 Jun 23;18(6):e0286297.

5. Ladds E, Rushforth A, Wieringa S, Taylor S, Rayner C, Husain L, et al. Developing services for long COVID: lessons from a study of wounded healers. Clin Med. 2021 Jan;21(1):59–65.

6. Kingstone T, Taylor AK, O’Donnell CA, Atherton H, Blane DN, Chew-Graham CA. Finding the ‘right’ GP: a qualitative study of the experiences of people with long-COVID. BJGP Open. 2020 Dec;4(5):bjgpopen20×101143.

7. Macpherson K, Cooper K, Harbour J, Mahal D, Miller C, Nairn M. Experiences of living with long COVID and of accessing healthcare services: a qualitative systematic review. BMJ Open. 2022 Jan;12(1):e050979.

8. Hossain MM, Das J, Rahman F, Nesa F, Hossain P, Islam AMK, et al. Living with “long COVID”: A systematic review and meta-synthesis of qualitative evidence. Canzan F, editor. PLoS ONE. 2023 Feb 16;18(2):e0281884.

9. Elliott P, Whitaker M, Tang D, Eales O, Steyn N, Bodinier B, et al. Design and Implementation of a National SARS-CoV-2 Monitoring Program in England: REACT-1 Study. Am J Public Health. 2023 May;113(5):545–54.

10. Ward H, Atchison C, Whitaker M, Davies B, Ashby D, Darzi A, et al. Design and Implementation of a National Program to Monitor the Prevalence of SARS-CoV-2 IgG Antibodies in England Using Self-Testing: The REACT-2 Study. Am J Public Health. 2023 Nov;113(11):1201–9.

11. Cooper E, Lound A, Atchison CJ, Whitaker M, Eccles C, Cooke GS, et al. Awareness and perceptions of Long COVID among people in the REACT programme: Early insights from a pilot interview study. Fauk NK, editor. PLoS ONE. 2023 Jan 26;18(1):e0280943.

12. Baz SA, Fang C, Carpentieri JD, Sheard L. ‘I don’t know what to do or where to go’. Experiences of accessing healthcare support from the perspectives of people living with Long Covid and healthcare professionals: A qualitative study in Bradford, UK. Health Expectations. 2023 Feb;26(1):542–54.

13. Atchison CJ, Davies B, Cooper E, Lound A, Whitaker M, Hampshire A, et al. Long-term health impacts of COVID-19 among 242,712 adults in England. Nat Commun. 2023 Oct 24;14:6588.

14. Cooper E, Lound A, Atchison CJ, Whitaker M, Eccles C, Cooke GS, et al. Awareness and perceptions of Long COVID among people in the REACT programme: Early insights from a pilot interview study. PLoS One. 2023;18(1):e0280943.

15. SAGE Publications Inc [Internet]. 2023 [cited 2023 Jul 11]. Thematic Analysis. Available from: https://us.sagepub.com/en-us/nam/thematic-analysis/book248481

16. Braun V, Clarke V. Thematic Analysis: A Practical Guide. SAGE; 2021. 377 p.

17. How public involvement changed our research question exploring experiences of people with Long Covid - Patient Experience Research Centre [Internet]. [cited 2024 Feb 14]. Available from: https://blogs.imperial.ac.uk/perc/2024/02/13/how-public-involvement-changed-our-research-question-exploring-experiences-of-people-with-long-covid/

18. Hossain MM, Das J, Rahman F, Nesa F, Hossain P, Islam AMK, et al. Living with “long COVID”: A systematic review and meta-synthesis of qualitative evidence. PLOS ONE. 2023 Feb 16;18(2):e0281884.

19. Soriano JB, Murthy S, Marshall JC, Relan P, Diaz JV. A clinical case definition of post-COVID-19 condition by a Delphi consensus. The Lancet Infectious Diseases [Internet]. 2021 Dec 21 [cited 2022 Mar 3]; Available from: https://www.sciencedirect.com/science/article/pii/S1473309921007039

20. Roth PH, Gadebusch-Bondio M. The contested meaning of “long COVID” – Patients, doctors, and the politics of subjective evidence. Social Science & Medicine. 2022 Jan 1;292:114619.

21. Ireson J, Taylor A, Richardson E, Greenfield B, Jones G. Exploring invisibility and epistemic injustice in Long Covid—A citizen science qualitative analysis of patient stories from an online Covid community. Health Expectations. 2022;25(4):1753–65.

22. Grue J. ILLNESS IS WORK: Revisiting the concept of illness careers and recognizing the identity work of patients with ME/CFS. Health (London). 2016 Jul;20(4):401–12.

23. Mengshoel AM, Skarbø Å, Hasselknippe E, Petterson T, Brandsar NL, Askmann E, et al. Enabling personal recovery from fibromyalgia – theoretical rationale, content and meaning of a person-centred, recovery-oriented programme. BMC Health Serv Res. 2021 Dec;21(1):339.

24. Dumit J. Illnesses you have to fight to get: Facts as forces in uncertain, emergent illnesses. Social Science & Medicine. 2006 Feb;62(3):577–90.

25. Marmot M. Health equity in England: the Marmot review 10 years on. BMJ. 2020 Feb 24;m693.

26. Siddiq S, Ahmed S, Akram I. Clinical outcomes following COVID-19 infection in ethnic minority groups in the UK: a systematic review and meta-analysis. Public Health. 2023 Sep;222:205–14.

27. Murali M, Gumber L, Jethwa H, Ganesh D, Hartmann-Boyce J, Sood H, et al. Ethnic minority representation in UK COVID-19 trials: systematic review and meta-analysis. BMC Med. 2023 Mar 29;21(1):111.

28. Corbin J, Strauss A. Managing chronic illness at home: Three lines of work. Qual Sociol. 1985;8(3):224–47.

29. Cooper H, Poland F, Kale S, Shakespeare T. Can a disability studies-medical sociology rapprochement help re-value the work disabled people do within their rehabilitation? Sociology Health & Illness. 2023 Jul;45(6):1300–16.

30. Bury M. Chronic illness as biographical disruption. Sociology of Health & Illness. 1982;4(2):167–82.

31. Rosland AM, Heisler M, Piette JD. The impact of family behaviors and communication patterns on chronic illness outcomes: a systematic review. J Behav Med. 2012 Apr;35(2):221–39.

32. Workers’ experience of Long Covid [Internet]. 2023 [cited 2024 Jan 3]. Available from: https://www.tuc.org.uk/research-analysis/reports/workers-experience-long-covid

33. Greene J, Hibbard JH. Why Does Patient Activation Matter? An Examination of the Relationships Between Patient Activation and Health-Related Outcomes. J Gen Intern Med. 2012 May;27(5):520–6.

