## Supplementary material for "Challenges of visibility and inclusion in Long COVID: a population-based interview study": Topic guide for interviews

**<u>S1 Topic Guide: Experiences of Covid</u>**

To explore the experiences and journeys of people with persistent symptoms of Covid-19

*<u>Remind participants to only answer questions they feel comfortable with</u>*

### Background

#### Can you tell me a little bit about your life before the pandemic?

- Living situation
- Employment status/field of work (prior to pandemic)
  - Key worker (eg NHS staff, teacher etc.)
- Activities
  - Sports
  - Hobbies
  - Travel
  - Things they do for fun
- Social life
  - Family
  - Friends
- General view of health and lifestyle
  - Sleep
  - Mental health
  - Exercise and diet

#### Can you tell me about your health history?

- Any pre-existing health conditions (ongoing)
- Any prior illnesses
  - Recovery time from illness in general
- Any prior unexplained/undiagnosed symptoms

### Experience of the pandemic

#### Can you give me a short summary of what the pandemic has been like for you?

- Living situation
- Employment
- Challenges

### Initial experience of COVID-19

#### I understand that you were diagnosed as having had COVID-19 at some point. Can you describe to me what your experience was??

- When did you suspect you had Covid-19? (*Probe if multiple times*)
- Did you have symptoms?
- Were you tested (within the first 5 days of symptoms)?
- Where/ how did you get tested?
- What symptoms did you experience?
    - Type (Cough, breathlessness, loss of taste or smell, fatigue, fever…)
    - Severity
- How did the illness progress?
    - Duration of initial illness
- What did you do?

#### [If had symptoms] What were your thoughts/feelings when you were first unwell?

- What did you know about COVID-19?
- What did you think was happening to you/your body?
- What did you think was going to happen to you/your body?
    - Hospitalisation
    - Death
    - Recovery
- How did you feel emotionally?

#### [If had symptoms] What help and/or support did you receive?

- Did you have anyone to look after you?
- Did you talk to anyone (friends/family) about your symptoms/how you felt?
- Can you tell me about any medical help you sought?
    - Medical advice: 111/pharmacy/GP/999/hospital/other
- Did you know other people (family, friends, colleagues) who were also affected?

### Ongoing journey

#### What happened after your initial illness?

- What happened next?
    - Persistent/changing/new symptoms after Covid-19
    - Initial recovery
    - General health changes

- Other physical or mental changes
  - Have you noticed any changes to your menstrual cycle or your periods following Covid? (Clarify that the participant does not have to answer this line of questioning)
  - Have you noticed any changes or your mood or mental health?
- What did you think was happening to you?
- What did you think was causing your symptoms?
  - Do you think anything about you/your situation had an effect on you developing these symptoms?
    - Has anything made your symptoms worse/better
- Did you talk to anyone or seek out any support for your ongoing symptoms?
  - Friends/family
  - Doctors (*probe on this if mentioned – see section 5*)
  - Support groups
- Can you describe the conversations you had

  How did you feel emotionally/mentally?
- How did things change?

##### Can you describe how different areas of your life were (or are) impacted?

- Can you describe to me to me what a 'good' or 'bad' day looks like
- Thinking about some of the examples of life before the pandemic we talked about at the start, can you tell me how things have changed?
  - Employment
  - Physical activities: walking short or long distances/climbing stairs/ standing/sitting
  - Daily life: Cooking/cleaning/personal care/entertainment
  - Activities: Work/education/socialising/exercise/hobbies
  - Family life and relationships
    - If you feel comfortable speaking about it could you tell me if anything has changed for you in terms of sex and intimacy? (*Clarify that the participant does not have to answer this line of questioning)*
- How does this make you feel?

#### Treatments and recovery

##### Can you tell me about any medical consultations, tests or treatments you sought or received? (*if not discussed above*)

- Did you find it easy or difficult to access help or treatment?

- Do you remember any particular conversations you had?
- Were you referred?
- How did you find talking to someone about your symptoms?
- Was this helpful?
- Did you receive a diagnosis?
    - How did that make you feel?

##### Can you tell me about anything you tried to help improve your symptoms?

- Alternative treatments
- Diet changes
- Lifestyle changes
- Supplements

Do you think anything in particular helped with your recovery?

##### What are your expectations/hopes for the future?

- What percentage back to normal are you now?
- Do you think you will recover?
    - What would recovery look like to you?
    - How long do you think recovery will take?
    - What do you think you need in order to recover?

Where on your recovery journey do you think you are now?

##### What are your thoughts on the COVID-19 vaccine?

- Have you had the vaccine?
- Which did you have, and when?
- Did you experience any side effects?
- Did you notice any changes to your symptoms?

#### Thoughts on Long COVID

##### Can you talk to be a bit about your general views on the term 'Long Covid'?

Are you aware of how 'Long Covid' was identified and where this term came from?

What do you think about the term 'Long Covid'?

- Does it cover what you are experiencing?
- Do you think this term is helpful/useful?

  - Do you think there should be another term (or terms) used instead?
  - What are your thoughts on the term ‘Post-Covid syndrome’
  - How do you think Long Covid is perceived by the public?
  - What others debates or discussions are you aware of around Long Covid**?**

#### Can you tell me about your thoughts on the diagnosis of persistent or new symptoms of Covid-19?

What does (or what would) a clinical diagnosis of your persistent or new symptoms mean to you?

Are you aware of any challenges around gaining a diagnosis?

What are your thoughts on the grouping of symptoms into multiple different diagnoses? (I.e symptoms clusters)

What do you think about persistent or new symptoms being compared to other illnesses e.g. ME/CFS

- In what ways do you think this is helpful/unhelpful?

#### What services do you think would be/ would have been helpful for you and others with persistent and new symptoms after Covid-19?

- Who should provide these services
  - GP/clinics/hospitals/community support
- What information would you like
- What wider support do you think should be available for people suffering with Long Covid
  - Functional/economic/physical/mental health/wellbeing

#### Can you tell me about any support groups or communities you may have joined?

- What groups did you join?
  - Online: Body politic slack/Long Covid Facebook group/twitter
  - Offline support: Community support/local groups
- What prompted you to join/not to join them?
- What sort of support/information do they provide?
- How do you feel about the group(s) now compared to when you first joined?

#### Wrap up

Is there anything we've not talked about that you think is important or that you would like us to know?

*Thank you for your time*
